# Diagnostic accuracy of a host response point-of-care test in patients with suspected COVID-19

**DOI:** 10.1101/2020.05.27.20114512

**Authors:** Tristan W Clark, Nathan J Brendish, Stephen Poole, Vasanth V Naidu, Christopher Mansbridge, Nicholas Norton, Helen Wheeler, Laura Presland, Sean Ewings

## Abstract

**Rationale:** Management of the COVID-19 pandemic is hampered by long delays associated with centralised laboratory PCR testing. In hospitals this leads to poor patient flow and nosocomial transmission and rapid, accurate diagnostic tests are urgently required. The FebriDx is a point-of-care test that detects an antiviral host response protein in finger prick blood within 10 minutes, but its accuracy for the detection of COVID-19 is unknown.

**Objectives:** To evaluate the diagnostic accuracy of FebriDx in hospitalised patients during the first wave of the pandemic

**Methods:** Measures of diagnostic accuracy were calculated based on FebriDx results compared to the reference standard of PCR, and stratified by duration of symptoms. A multivariable predictive model was developed and underwent internal validation.

**Results:** FebriDx was performed on 251 patients and gave a valid result in 248. 118 of 248 (48%) were PCR positive for COVID-19. Sensitivity of FebriDx for the identification of COVID-19 was 93% (110/118; 95% CI 87 to 97%) and specificity was 86% (112/130; 95%CI 79 to 92%). Positive and negative likelihood ratios were 6.73 (95%CI 4.37 to 10.37) and 0.08 (95%CI 0.04 to 0.15) respectively. In the multivariate model diagnosis of COVID-19 was not significantly influenced by clinical symptoms and signs, and FebriDx accuracy was not improved by restricting testing to those with duration of symptoms of less than seven days.

**Conclusions:** During the first wave of the pandemic, FebriDx had high sensitivity for the identification of COVID-19 in hospitalised adults and could be deployed as a front door triage tool.

**Trial registration:** ISRCTN14966673

## Introduction

The management of COVID-19 cases in secondary care is severely hampered by the long turnaround times of centralised laboratory PCR testing, which can take up to several days to generate results. In acute hospitals this leads to poor patient flow though acute areas, as suspected patients are held in assessment cohort areas until their results are available. In addition, lack of single occupancy rooms means that COVID-19 negative patients cohorted in these assessment areas may acquire infection from positive patients before test results are available. Rapid, accurate diagnostic tests are therefore urgently required. Molecular point-of-care testing (POCT) for COVID-19 may mitigate this situation but regulatory requirements for new POCT assays and difficulties in rapidly upscaling manufacture mean that the availability of these tests is currently severely limited. Alternative diagnostic solutions are therefore urgently required.

FebriDx (Lumos diagnostics, Sarasota, Florida, US) is a CE-marked POCT that detects two host response proteins, Myxovirus resistance protein A (MxA) and C reactive protein (CRP), in finger prick blood samples, and is designed to distinguish viral from bacterial respiratory infection.^1-5^ MxA is a marker of interferon-induced antiviral host response and in our previous work, the detection of MxA by FebriDx had high sensitivity for the identification of Influenza in hospitalized adults, during influenza season.^6^ MxA levels are also likely to be significantly elevated in patients with COVID-19 but the diagnostic accuracy of FebriDx in this situation is currently unknown. FebriDx is a low cost, instrument-free, disposable POCT system and if sensitive for the detection of COVID-19 could be rapidly deployed across the NHS and other healthcare systems as a rapid triage tool, and help mitigate the long delays asdociated with current diagnostic testing. The aim of this study was therefore to evaluate the real-world diagnostic accuracy of MxA detection by FebriDx for the identification of COVID-19 in hospitalised adults during the first wave of the pandemic, and to develop and internally validate a multivariable model for diagnosis of COVID-19 that includes FebriDx result and other easily obtained clinical measures.

## Methods

### Study design and participants

This study of FebriDx was nested within the CoV-19POC study, a trial assessing the clinical impact of molecular POCT for COVID-19. Adults presenting with suspected COVID-19 were enrolled; full details of the inclusion and exclusion criteria can be found in the protocol, linked below. The study was approved by the South Central - Hampshire A Research Ethics Committee: REC reference 20/SC/0138, on the 16th March 2020. This trial is ongoing and the protocol is available at: https://eprints.soton.ac.uk/439309/1/CoV_19POC_Protocol_v1.1_eprints.pdf

### Procedures

Combined nose and throat swabs were obtained from patients by research staff and tested immediately on the QIAstat-Dx platform using the Respiratory SARS-CoV-2 Panel,^7^ at the point of care. A full list of the pathogens detected by the panel is shown in Table E1 in the online data supplement.^8, 9^ In addition, laboratory PCR testing for SARS-CoV-2 on combined nose and throat swabs was performed on all patients, in the on-site Public Health England (PHE) microbiology laboratory, using the PHE RdRp and E gene reference assays.^10, 11^ Demographic and clinical data was collected at enrolment and outcome data collected retrospectively from case note and electronic systems, using an electronic case report form.

For this sub-study the first 266 patients enrolled were consecutively approached for testing using the FebriDx host response POCT on finger prick blood samples, taken at the same time as the nose as throat swabs for PCR. Detailed instructions for use of FebriDx are provided via this link: https://www.febridx.com/how-to-use#testing but in brief; after puncture of the skin with the integral lancet, 5 microliters of blood are drawn into the blood collection tube via capillary action by placing it against a blood drop. The blood is then transferred to the lateral flow section of the device and reagents released by pressing a button. The test is read after 10 minutes.

### Interpretation of results

The FebriDx system generates results in the form of the presence or absence of three lines, assessed by visual inspection; the CRP line (top, grey), the MxA line (middle, red) and the control line (bottom, blue). Figure 2 shows examples of FebriDx positive and negative tests. The colour change on the CRP and MxA lines when detected is dependent on the amount of the target protein in the sample and so there is potential variability in interpretation with weaker lines (quoted threshold for detection of CRP line 20mg/L and MxA line of 40ng/ml). For this reason each FebriDx result was read independently by two investigators and if there was disagreement on the results displayed this was further adjudicated by a third investigator. The results of the FebriDx were not shared with the clinical teams and the readers of the FebriDx test lines were blinded to the PCR results (FebriDx results were read and recorded before PCR results were available).

For the reference standard of PCR, the QIAstat-Dx PCR system gives a readout of positive or negative for the detection of targets including SARS-CoV-2.^7^ The PHE laboratory RdRp and E gene assay PCR results are also provided as a binary result, RNA detected or RNA not detected, and the detection of RNA by either assay is considered a positive result. For this study PCR detection by either QIAstat-Dx or laboratory PCR or both was considered as positive for COVID-19.

### Sample size

The sample size for this sub-study was driven by consideration for estimating sensitivity of FebriDx MxA for the identification of COVID-19 (as defined by the reference standard of detection of SARS-CoV-2 by PCR on respiratory samples) and of the methods proposed by Riley et al^12^ for multivariable predictive models. In order to estimate a sensitivity of 85% to within +/- 8%, based on the score method for a 95% confidence interval,^13^ 88 positive cases are required. Assuming a prevalence of COVID-19 of around 40% in those tested, 220 people are required, and to achieve an 80% chance of obtaining enough cases, this number was increased to 236. This was further increased to 266 to allow for patients who decline or are unable to undergo finger prick testing (estimated from the rates in our previous work). Full justification for the samples size in the multivariable model is provided in File E1 in the online data supplement.

### Statistical analysis

Baseline characteristics are summarised for all those recruited to the study, where data are available, and presented for the whole sample and by COVID-19 status. Diagnostic accuracy was assessed in two ways. Simple measures of sensitivity, specificity, predictive values and likelihood ratios are given for FebriDx MxA detection for the identification of COVID-19 (and as FebriDx MxA is a broad marker of antiviral host response and not limited to COVID-19 these measures are also calculated for the detection of all viruses). To further assess the diagnostic value of the test in practice, a multivariable logistic regression model was developed,^14,15^ full details are provided in file E1 in the online data supplement.

As the antiviral host response to SARS-CoV-2 would be expected to wane over time, but detection of viral RNA by PCR may persist for several weeks, a secondary analysis was pre-specified to assess diagnostic accuracy in those who have had duration of less than seven days. Basic measures of diagnostic accuracy (e.g., sensitivity, specificity) are presented separately for those with duration less than or more than seven days. A multivariable model, pre-specified to be the same as that for the full model, was also run in those with duration less than seven days.

This study was prospectively registered with the ISRCTN14966673 on the 18^th^ March 2020.

## Results

The first 266 patients recruited to the CoV-19POC study were enrolled and FebriDx was performed in 251. 12 of 251 (5%) tests failed to give a valid result on the first test and a subsequent valid result was obtained on re-testing in 9 of 12, so that overall 248 patients had a valid FebriDx result. Flow of study participants is shown in Figure 1. 118 of 248 (48%) were COVID-19 positive by PCR (for SARS-CoV-2) on combined nose and throat swabs and 92 of 116 (79%) COVID-19 positive patients had pneumonia as defined by Chest X-ray changes. PCR and FebriDx results were available for all tested patients. For symptoms, 8 to 13% of data was missing and for vital signs and demographics missing data was <2.5%. Results are presented using complete-case analysis. Results including anosmia are included as an additional secondary analysis. Data collection for this was initiated after recruitment started, once it came to light in other literature that this was a relevant symptom of COVID-19; a total of 64 of 248 (26%) patients had missing data. Baseline demographics and clinical characteristic of all patients and those testing positive and negative for COVID-19, are shown in Table 1. Patients positive for COVID-19 were less likely to be ethnically White British, had been unwell for longer (median duration of illness prior to presentation of 8 days), had a higher temperature and respiratory rate and lower oxygen saturations, compared to COVID-19 negative patients. In addition, COVID-19 positive patients had higher CRP levels, lower white cell counts and were more likely to have radiological evidence of pneumonia, shown in Table 1.

**Figure 1.**
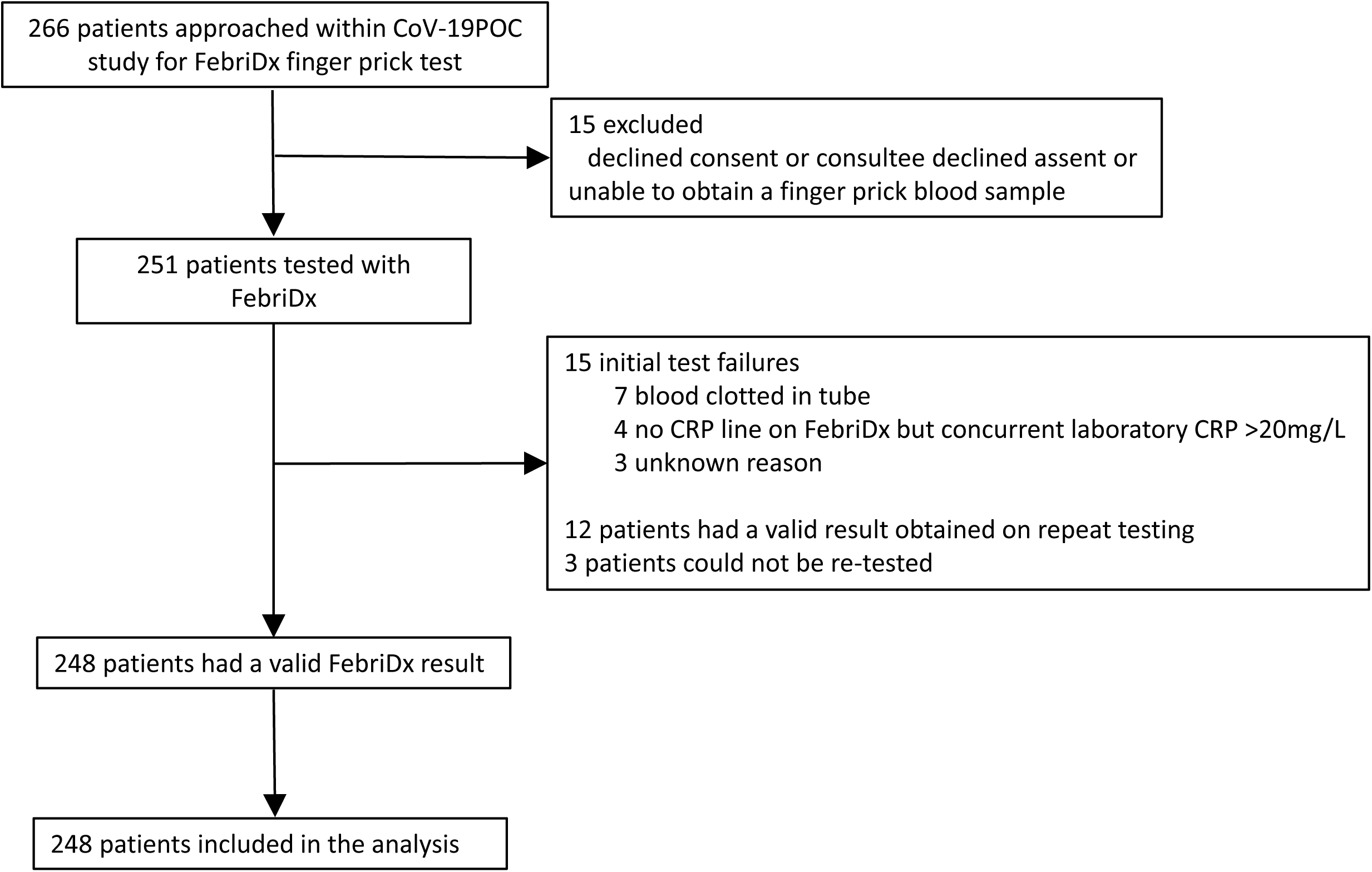
Flow of participants through the study

**Figure 2.**
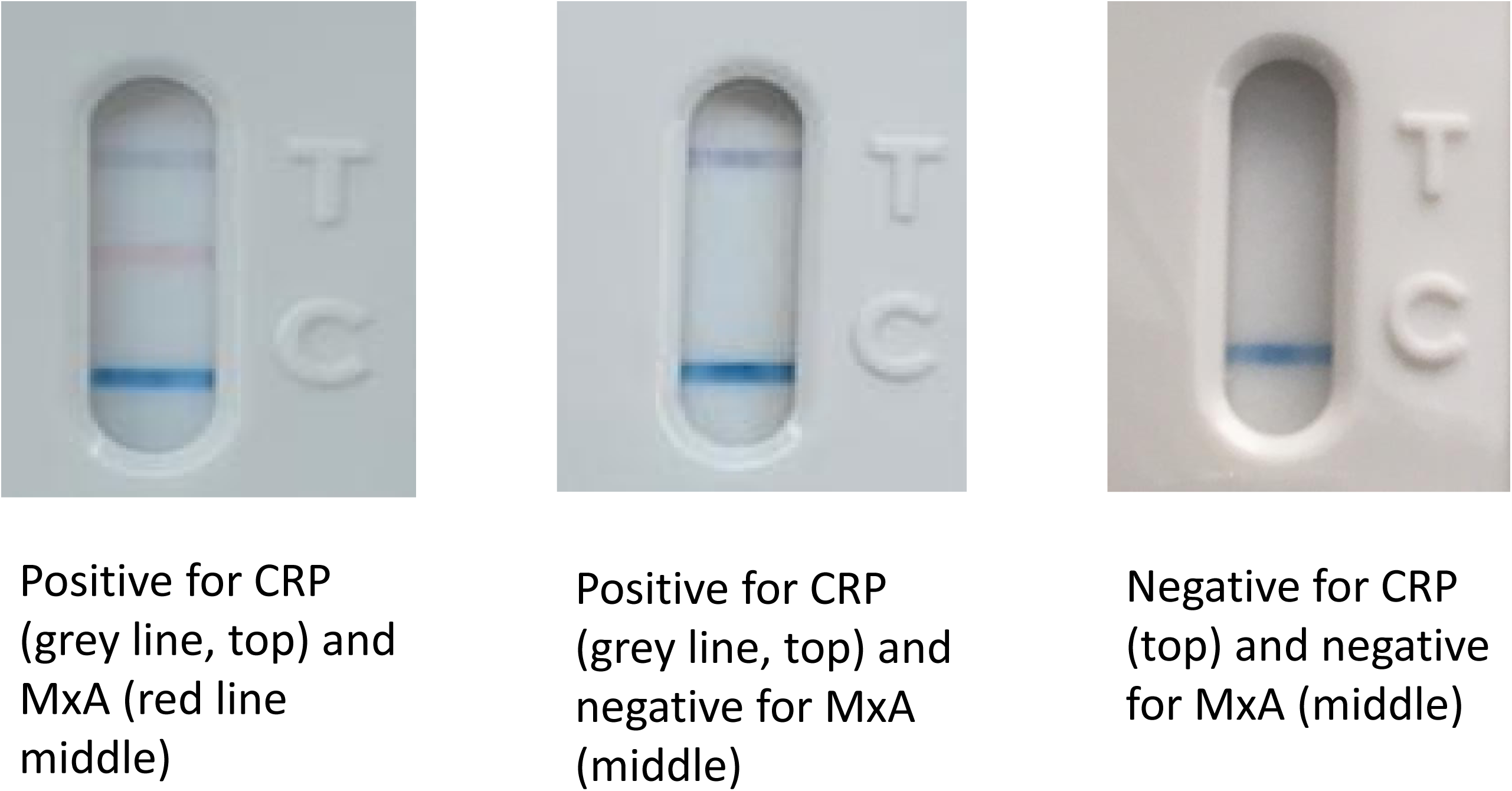
Examples of positive and negative FebriDx read-outs

**Table 1.** Baseline demographics and clinical characteristics in all patients, COVID-19 positive patients and COVID-19 negative patients

|  | **All patients**<br>**n=248** | **COVID-19 positive**<br>**patients**<br>**n=118** | **COVID-19 negative**<br>**patients**<br>**n=130** | **p value\*** |
| --- | --- | --- | --- | --- |
| Age, years | 70 (52 to 81) | 67 (49 to 79) | 72 (58 to 81) | 0.13 |
| Sex |  |  |  |  |
| Male | 135/248 (54) | 69/118 (58) | 66/130 (51) | 0.25 |
| Female | 113/248 (46) | 49/118 (42) | 64/130 (49) | - |
| White British | 219/245 (89) | 94/116 (81) | 125/129 (97) | <0.0001 |
| Current smoker | 21/211 (10) | 6/102 (6) | 15/109 (14) | 0.067 |
| Influenza vaccine† | 127/189 (67) | 63/92 (69) | 63/97 (65) | 0.53 |
| Pregnant | 4/46 (9) | 2/22 (9) | 2/25 (8) | 0.99 |
| Duration of symptoms, days‡ | 3 (1 to 7) | 8 (5 to 12) | 4 (2 to 11) | 0.0006 |
| **Comorbidity** |  |  |  |  |
| Hypertension | 97/246 (39) | 48/117 (41) | 49/129 (38) | 0.69 |
| Cardiovascular disease | 79/247 (32) | 35/118 (30) | 44/129 (34) | 0.49 |
| Respiratory disease | 98/248 (40) | 37/118 (31) | 61/130 (47) | 0.014 |
| Renal disease | 20/245 (8) | 8/118 (7) | 12/127 (9) | 0.49 |
| Liver disease | 8/247 (3) | 4/118 (3) | 4/129 (3) | 1.0 |
| Diabetes mellitus | 53/247 (22) | 25/118 (21) | 28/129 (22) | 0.88 |
| Cancer | 16/247 (7) | 8/118 (7) | 8/129 (6) | 1.0 |
| Immunosuppression | 10/244 (4) | 5/117 (4) | 5/127 (4) | 1.0 |
| Dementia | 26/148 (11) | 9/118 (8) | 17/130 (13) | 0.21 |
| **Observations at admission** |  |  |  |  |
| Temperature, °C | 36.9 (36.5 to 37.8) | 37.2 (36.6 to 38.2) | 36.7 (36.4 to 37.4) | 0.0042 |
| Temperature ≥38°C | 55/242 (23) | 36/114 (32) | 19/128 (15) | 0.0021 |
| Pulse rate, bpm | 92 (81 to 108) | 92 (92 to 109) | 92 (81 to 108) | 0.91 |
| Respiratory rate, bpm | 24 (20 to 28) | 25 (20 to 28) | 23 (19 to 28) | 0.023 |
| Oxygen saturations, % | 96 (94 to 97) | 95 (92 to 97) | 96 (94 to 98) | 0.030 |
| Supplementary O<sub>2</sub> | 90/246 (37) | 45/116 (39) | 44/128 (34) | 0.51 |
| Blood pressure, mmHg |  |  |  |  |
| Systolic | 130 (120 to 146) | 130 (120 to 145) | 132 (118 to 150) | 0.52 |
| Diastolic | 75 (66 to 83) | 75 (68 to 83) | 74 (65 to 84) | 0.60 |
| NEWS2 score | 5 (3 to 6) | 5 (3 to 7) | 4 (2 to 6) | 0.017 |
| **Laboratory and radiology at admission** |  |  |  |  |
| CRP, mg/L | 60 (14 to 128) | 83 (32 to 136) | 33 (9 to 114) | 0.0003 |
| WCC, 10<sup>9</sup>/L | 8.5 (6.1 to 12.7) | 6.6 (5.0 to 8.7) | 11.1 (7.9 to 15.4) | <0.0001 |
| CXR performed | 241/245 (98) | 116/116 (100) | 125/129 (97) | 0.12 |
| **Final Diagnosis** |  |  |  |  |
| Pneumonia | 143 (58) | 92/116 (79) | 51/129 (40) | <0.0001 |
| Other | 102 (42) | 24/116 (21) | 78/129 (60) | - |

All data are given as n/n (%) and median (inter-quartile range). COVID-19 positive and negative status is defined by PCR positivity or negativity for SARS-CoV-2. \*P value relate to comparison between COVID-19 positive and negative patients. NEWS=National Early Warning Score 2, CRP=C reactive protein, WCC=White cell count. †For the most recent influenza season. ‡Duration of symptoms prior to presentation, evaluated in 215 patients.

### Diagnostic accuracy

Compared to the reference standard of detection of SARS-CoV-2 RNA by PCR on respiratory samples, the sensitivity of FebriDx MxA detection for the identification of COVID-19 was 93% (110/118; 95%CI 87 to 97%). Specificity in this cohort was 86% (112/130; 95%CI 79 to 92%). Positive predictive value (PPV) was 86% (110/128; 95% CI 79 to 91%) and negative predictive value (NPV) was 93% (112/120; 95% CI 87 to 97%). Overall accuracy was 90% (222/248; 95%CI 86 to 93%), shown in Table 2. As detection of MxA is not specific for COVID-19 the diagnostic accuracy of FebriDx MxA detection for the identification of all respiratory viruses is shown in Table 3. Of the 11 patients with positive FebriDx MxA but who were PCR negative for SARS-CoV-2 and other viruses, 7 of 11 (63%) had classic radiological features of COVID-19.

**Table 2.** Measures of diagnostic accuracy of FebriDx MxA for identification of COVID-19, compared to the reference standard of PCR positivity, n=248.

|  | **n/n** | **Value (95%CI)** |
| --- | --- | --- |
| Prevalence of Covid-19 | 118/248 | 48 (41 to 54) |
| Sensitivity | 110/118 | 93 (87 to 97) |
| Specificity | 112/130 | 86 (79 to 92) |
| Positive predictive value | 110/128 | 86 (79 to 91) |
| Negative predictive value | 112/120 | 93 (87 to 97) |
| Positive likelihood ratio |  | 6.73 (4.37 to 10.37) |
| Negative likelihood ratio |  | 0.08 (0.04 to 0.15) |
| Overall accuracy | 222/248 | 90 (86 to 93) |

CI=Confidence interval

**Table 3.** Measures of diagnostic accuracy of FebriDx MxA detection for any respiratory virus, compared to the reference standard of PCR positivity, n=248.

|  | **n/n** | **Value (95%CI)** |
| --- | --- | --- |
| Prevalence of any virus* | 135/248 | 54 (48 to 61) |
| SARS-CoV-2 (COVID-19) | 118/248 | 48 (41 to 54) |
| Rhino/enterovirus | 10/248 | 4 (2 to 7) |
| Human Metapneumovirus | 4/248 | 2 (1 to 4) |
| Human Coronavirus OC43 | 2/248 | 2 (1 to 4) |
| Human Coronavirus HKU1 | 1/248 | 1 (0 to 2) |
| Human Coronavirus NL63 | 1/248 | 1 (0 to 2) |
| Sensitivity† | 117/135 | 87 (80 to 92) |
| Specificity | 102/113 | 90 (83 to 95) |
| Positive predictive value | 117/128 | 91 (86 to 94) |
| Negative predictive value | 112/120 | 85 (78 to 90) |
| Positive likelihood ratio |  | 8.9 (5.1 to 15.7) |
| Negative likelihood ratio |  | 0.15 (0.1 to 0.23) |
| Overall accuracy | 219/248 | 88 (83 to 92) |

CI=Confidence interval. \*One patient had co-detection of SARS-CoV-2 and HCoV HKU1. † Viral detections associated with false negative MxA were: SARS-CoV-2 x 8, Rhino/enterovirus x 7 and Human Coronavirus x 3.

Using the calculated performance of FebriDx MxA for the identification of COVID-19 to estimate PPV and NPV at different disease prevalence, a prevalence of COVID-19 of 20% would give a PPV of 63% (95%CI 52 to 72%) and NPV of 99% (95%CI 96 to 99%). At a prevalence of 10% the PPV would be 43% (95%CI 33 to 53%) and the NPV would be 99% (95%CI 98 to 100%).

Measures of diagnostic accuracy according to duration of symptoms are shown in Table 4. Although direct comparison of accuracy was not possible due to samples size, the sensitivity, specificity and positive and negative predictive values of FebriDx appeared comparable for patients presenting before and after 7 days of symptom duration.

**Table 4.** Measures of diagnostic accuracy of FebriDx MxA for identification of COVID-19, according to duration of illness.

|  | **n/n** | **≤7 days of symptoms**<br>**n=129**<br>**Value (95% CI)** | **n/n** | **>7 days of symptoms**<br>**n=86**<br>**Value (95% CI)** |
| --- | --- | --- | --- | --- |
| Median (IQR) duration, days |  | 4 (1 to 6) |  | 14 (8 to 14) |
| Prevalence of COVID-19 | 52/129 | 40 (32 to 49) | 53/86 | 62 (51 to 71) |
| Sensitivity | 49/52 | 94 (84 to 98) | 48/53 | 91 (80 to 96) |
| Specificity | 65/77 | 84 (75 to 91) | 29/33 | 88 (73 to 95) |
| Positive predictive value | 49/61 | 80 (69 to 88) | 48/52 | 92 (82 to 97) |
| Negative predictive value | 65/68 | 96 (88 to 99) | 29/34 | 85 (71 to 93) |
| Positive likelihood ratio |  | 6.05 (3.58 to 10.0) |  | 7.47 (2.96 to 19.0) |
| Negative likelihood ratio |  | 0.07 (0.02 to 0.21) |  | 0.11 (0.5 to 0.25) |

IQR=Interquartile range, CI=Confidence interval

### Predictive model

A total of 201 participants contributed data to the multivariable model, with symptoms being the main contributor to missing data. With the possible exception of cough (12% missing data in those without COVID-19 versus 6% with), missingness of data appeared unrelated to outcome. All continuous variables (age, respiratory rate and temperature) were mean-centred and assessed for a non-linear relationship with outcome using restricted cubic splines, following visual assessment and using Bayesian Information Criterion values (where smaller values indicate a more parsimonious model), all were entered in the model on the assumption that a linear relationship with the log odds of having COVID-19 was appropriate. All variables were entered into the multivariable model with no variable selection. Model results are given in Table 5, alongside adjusted estimates.

**Table 5.** Results for multivariable model

| **Variable** | **Regression**<br>**coefficient**<br>**(odds ratio)** | **95%**<br>**confidence**<br>**interval** | **p-value** | **Adjusted**<br>**coefficient**<br>**(odds ratio)** |
| --- | --- | --- | --- | --- |
| FebriDx MxA result | 92.3 | 31.6 to 269.6 | <0.0001 | 49.8 |
| Cough | 2.1 | 0.67 to 6.9 | 0.21 | 1.9 |
| Fever | 0.9 | 0.3 to 2.9 | 0.85 | 0.9 |
| Shortness of breath | 0.7 | 0.2 to 2.1 | 0.48 | 0.7 |
| Supplementary O<sub>2</sub> | 0.7 | 0.3 to 1.9 | 0.49 | 0.7 |
| Sex | 1.0 | 0.4 to 2.7 | 0.92 | 1.0 |
| Age | 1.0 | 1.0 to 1.0 | 0.74 | 1.0 |
| Respiratory rate | 1.1 | 1.0 to 1.2 | 0.03 | 1.1 |
| Temperature | 1.2 | 0.7 to 1.9 | 0.52 | 1.1 |
| Intercept | 0.1 | 0.02 to 0.27 | <0.0001 | 0.1 |

Nagelkerke's R<sup>2</sup> = 0.71; Brier score 0.09. Unless otherwise indicated, all reference categories for categorical variables are no/absence.

The overall results suggested that the addition of the patient and clinical characteristics do not add significantly to the diagnostic accuracy compared to using FebriDx MxA result alone. The apparent area under the receiver-operating characteristic (ROC) curve (AUC; equivalent to the C-statistic) for the multivariable model was 0.93, suggesting very good discrimination between those with and without PCR-confirmed COVID-19. The calibration plot shows good calibration (Figure E1 in the online data supplement), with points lying near the 45-degree line representing perfect fit.

The model underwent internal validation, using bootstrap resampling with 500 samples following the same model development process, to estimate optimism and provide a basis for adjusting model coefficients and the C-statistic. The calibration slope suggests the model tended to estimate probabilities that are too extreme and that if the model were to be used externally, the original multivariable regression coefficients should be adjusted by the value of the calibration slope (the shrinkage factor) to adjust for optimism. The C-statistic was also adjusted for optimism, but remained very high (0.90) suggesting the model would still perform well outside of the current sample.

### Secondary analyses

The multivariable model was rerun with anosmia; a total of n=164 were included in this analysis. Conclusions for the other covariates did not change, and there was little change in discrimination results (area under the curve, sensitivity and specificity at the Youden index, Brier score) nor other measures of fit such as Nagelkerke’s R^2^; anosmia itself had an estimated odds ratio of 4.0 (95% CI: 0.7 to 22; p=0.11) suggesting a potentially useful diagnostic indicator, though this conclusion is hampered by the uncertainty in the estimate. Lastly, the multivariable model (excluding anosmia) was also run in the subgroup of participants who presented within 7 days of symptom onset. Although the odds ratio for FebriDx increased (190), the uncertainty of this effect was large (95% CI 26 to 1404). There was little change in model performance with regard discrimination and model fit.

### Test failures

As detailed above, some FebriDx tests failed to give a valid result on the first attempt. Tests could fail by blood clotting in the sample collection tube so that no blood would pass onto the lateral flow strip.

### Safety

There were no adverse events seen with FebriDx testing. A single patient suffered brief epistaxis following nasal swabbing for PCR testing.

## Discussion

In this large, real-world study we have shown that that the detection of the antiviral host response protein MxA using the FebriDx point-of-care test had high sensitivity for the identification of COVID-19 during the first wave of the pandemic. Although there were differences in baseline characteristics between COVID-19 positive and negative patients, the multivariate predictive model results showed that clinical features did not contribute significantly to the effect of the FebriDx MxA result in distinguishing these groups. MxA detection is a marker of antiviral host response and is not specific for SARS-CoV-2 and so the high specificity of FebriDx MxA for COVID-19 seen in this study reflects the low levels of other respiratory viruses circulating during the study which will change with the seasonal circulation of other viruses such as influenza. It is now recognised that the diagnostic accuracy of PCR on upper respiratory tract samples is suboptimal and that patients presenting with COVID-19 often initially test falsely negative when compared to serial PCR testing or CT scanning.^16-18^ This means that our reference standard was likely to be suboptimal and it is notable that several of the patients in our study with positive MxA results and negative PCR for viruses (ie those considered to be false positive MxA) had classical radiological changes of COVID-19, strongly suggesting that they had the disease despite negative PCR results. The calculated specificity of FebriDx MxA for the detection of COVID-19 in the first wave was therefore likely to have been an underestimate.

The current reliance on centralised laboratory PCR for the diagnosis of COVID-19 with its prolonged turnaround time, leads to long delays in identification of positive and negative patients, with subsequent nosocomial transmissions, poor patient flow and reduced operational capacity in hospitals. As FebriDx is a low cost (£12 per test), rapid and highly scalable test these findings suggest that FebriDx could be rapidly deployed in hospitals and urgent care centres to be used as a front door triage tool. Because of the high sensitivity and negative predictive value, MxA negative patients being admitted to hospitals could be rapidly directed to non COVID-19 areas whilst MxA positive patients could be moved to assessment areas whilst awaiting confirmation or exclusion of COVID-19 positive status by PCR or CT scanning.

The manufacturers of FebriDx state that it is intended for use in patients aged over two years of age who have had respiratory symptoms for less than or equal to 7 days and/or fever for 48 hours duration or less. In this study we have demonstrated that MxA detection by FebriDx remains accurate for the identification of COVID-19 beyond this period, presumably due to a very strong and persistent antiviral host response to SARS-CoV-2.

There are several limitations to the generalisability of our study. As the study was performed in almost exclusively immunocompetent adults the results cannot be applied to immune-compromised patients or to children and separate evaluations should be urgently undertaken in these important patient groups. In addition the findings of our study cannot be extrapolated to community dwelling patients including those who are infected but asymptomatic or pauci-symptomatic, as it uncertain whether their antiviral host response would be comparable to hospitalised patients. The diagnostic accuracy of FebriDx reported in this study is in the context of a very high disease prevalence and as we have shown the performance characteristics will change as the disease prevalence drops, with implications for how the test could be used. In particular the positive predictive value will decrease now that the first wave is subsiding, although the negative predictive value will increase further meaning that a negative FebriDx will remain a useful rule-out test. As MxA detection is a marker of antiviral host response and is not specific for COVID-19, the calculated test specificity will be lower when other viruses such as influenza are circulating. As early identification of influenza alongside COVID-19 will remains vitally important in hospitals, a rapid triage tool that detects both influenza and COVID-19 is likely to be of utility in the coming winter months. Finally, as this was a single centre study the diagnostic accuracy of FebriDx MxA for COVID-19 should be external validated in other centres.

In summary, detection of MxA by FebriDx had a high sensitivity for the identification of COVID-19 in the context of a high prevalence during the first wave of the pandemic. FebriDx could be rapidly deployed in secondary care settings as a triage tool to address the current problems of delayed diagnosis with laboratory PCR.

## Data Availability

Data requests should be submitted to the corresponding author for consideration. Following publication access to anonymised data may be granted following review

## Acknowledgements

We would like to acknowledge and gives thanks to all the patient who kindly participated in this study and to all the clinical staff at University Hospital Southampton who cared for them. We would also like to give thanks to Richard Poole for assistance in data management.

This report is independent research supported by the National Institute for Health Research (**NIHR Post Doctorial Fellowship, Dr Tristan Clark, PDF 2016-09-061**).

The views expressed in this publication are those of the author(s) and not necessarily those of the NHS, the National Institute for Health Research or the Department of Health.

## Online Data Supplement

**Table E1**. Pathogens tested for by QIAstat-Dx Respiratory SARS-CoV-2 Panel

**File E1**. Sample size justification and methods for multivariable predictive model

**Figure E1**. Calibration plot for multivariate model

